# Using risk advancement periods to derive starting ages of colorectal cancer screening according to sex and polygenic risk score: Results from the UK Biobank

**DOI:** 10.1101/2023.03.05.23286808

**Authors:** Xuechen Chen, Thomas Heisser, Rafael Cardoso, Michael Hoffmeister, Hermann Brenner

**Affiliations:** Division of Clinical Epidemiology and Aging Research, German Cancer Research Center (DKFZ), Heidelberg 69120, Germany; Medical Faculty Heidelberg, Heidelberg University, Heidelberg 69120, Germany; Division of Preventive Oncology, German Cancer Research Center (DKFZ) and National Center for Tumor Diseases (NCT), Heidelberg 69120, Germany; German Cancer Consortium (DKTK), German Cancer Research Center (DKFZ), Heidelberg 69120, Germany

## Abstract

**Objective:** Polygenic risk scores (PRSs) derived from genome-wide association studies are strong predictors of colorectal cancer (CRC) risk. We applied the straightforward approach of risk advancement periods (RAPs) to derive risk-adapted starting ages of CRC screening according to sex and PRS in the UK Biobank.

**Methods:** Among 242,779 participants (40-69 years; no previous CRC screening; no family history of CRC), we assessed associations of sex and a PRS with CRC risk and mortality using Cox regression models. Hazard ratios (HRs) were translated to RAPs to quantify how many years of age earlier men and women in defined PRS deciles reach comparable risks as those in the reference group (5^th^ and 6^th^ PRS deciles).

**Results:** During a median follow-up of 11.2 and 12.8 years, 2,714 participants were diagnosed with CRC and 758 died from CRC, respectively. HRs (95% CIs) of CRC risk were 1.57 (1.46, 1.70) for men versus women and ranged from 0.51 (0.41, 0.62) to 2.29 (2.01, 2.62) across PRS deciles compared to the reference. RAPs (95% CI) were 5.6 (4.6, 6.6) years for men versus women, and ranged from -8.4 (-11.0, -5.9) to 10.3 (8.5, 12.1) years across PRS deciles compared to the reference. Risk-adapted starting ages would vary by 24 years between men in the highest PRS decile and women in the lowest PRS decile. Very similar results were obtained regarding CRC mortality.

**Conclusion:** Consideration of sex and a standard PRS alone could have far-reaching implications for starting ages of CRC screening in the “average risk population”.

**SUMMARY BOX:** *What is already known on this topic:*

- Polygenic risk scores (PRSs) are strong predictors of colorectal cancer (CRC) risk.
- Men have substantially higher CRC incidence and mortality than women.
- CRC risk information from the combination of PRS and genetically determined sex, which are constant factors over lifetime, is so far not used for risk-adapted CRC screening in the “average risk population”.

*What this study adds:*

- Risk advancement periods (RAPs) of CRC by PRS and sex were derived at high levels of precision from the large database of the UK Biobank.
- By joint consideration of PRS and genetically defined sex, risk-adapted starting ages would vary by as much as 24 years between men in the highest PRS decile and women in the lowest PRS decile.

*How this study might affect research, practice or policy:*

- Our study demonstrates a straightforward way to translate CRC risk information from a large population-based cohort into risk-adapted starting ages of screening.
- Personalized, risk-adapted starting ages of CRC screening could be derived from a single blood test performed in middle adulthood.
- The RAP approach could be easily extended for defining personalized starting ages by incorporating additional risk factors in the regression models.

## Introduction

Colorectal cancer (CRC) is still the second most common cause of cancer-related death globally [1], even though there are effective ways of early detection that could substantially reduce the burden of the disease [2,3]. Screening for CRC is meanwhile recommended and offered to the average risk population in many countries, but screening recommendations, offers and use vary substantially between countries [4,5]. For example, CRC screening, either by screening colonoscopy, fecal immunochemical test or a DNA-based stool test is now recommended starting from age 45 on in the United States [6], whereas screening programs starting at ages between 40 and 60 years have been implemented, or even no screening is offered at all, in various European countries [5]. Besides variation in CRC incidence between countries, there is increasing evidence for substantial variation of CRC risk within populations, which has prompted suggestions for risk-adapted, personalized screening offers [7–10]. A particularly relevant issue in this respect is if and to what extent starting ages for CRC screening should be adapted to personal risks. Although recommendations for earlier screening among people with first-degree relatives of CRC have long been established [11,12], other major risk determinants are not routinely considered in established screening recommendations.

Generally, two types of risk determinants might be relevant in this context: risk determinants that are “fixed” throughout lifetime, and modifiable risk determinants, such as lifestyle habits, that may change over lifetime. Fixed determinants may be particularly relevant for determining starting ages of screening, as they are expected to exert their impact from young ages on. Modifiable risk determinants would be most relevant for efforts of primary prevention and may change over time, accumulate their effects throughout adulthood and might be increasingly relevant at older ages. Even family history of CRC risk, a non-modifiable risk factor, often becomes evident at older ages only, e.g., beyond commonly recommended starting ages of screening [9,13]. This particularly applies to CRC among siblings, as mean age at CRC diagnosis is around 70 years or older in many high-income countries.

Two major CRC risk factors that are fixed throughout lifetime and that could be reliably determined already in young or middle adulthood, i.e., at pre-CRC screening ages, are (genetically determined) sex and genetically determined CRC risk variants, as reflected in polygenic risk scores (PRSs) [14,15]. In this study, we applied the straightforward concept of risk advancement periods [16] to derive risk-adapted starting ages of screening at which comparable levels of CRC occurrence or mortality risk are reached according to sex and PRS in a large European cohort.

## Methods

### Study Design and Study Population

This research has been conducted using the UK Biobank Resource under Application Number 66591. The UK Biobank study is a population-based cohort study with over half a million adults recruited at ages 40-69 years in 2006-2010 across England, Scotland and Wales. In addition to the collection of biological samples (blood, saliva and urine), information on sociodemographic, health and medical history, anthropometric and lifestyle factors was collected in one of 22 UK assessment facilities. Follow-up of health-related outcomes is conducted through linkage to electronic health records including death, cancer, primary care and hospital admissions from the UK National Health Service. Further details of the UK Biobank study protocol have been published elsewhere [17]. The UK Biobank study has obtained approval from the North West Multi-center Research Ethics Committee (MREC) as a Research Tissue Bank (RTB) approval (renewed approval in 2021: 21/NW/0157). All participants provided electronic signed informed consent.

### Genotyping and Imputation

Details of genotyping, quality control and imputation have been published elsewhere [18,19]. Briefly, genotyping was done using two arrays, the UK BiLEVE Axiom (∼50,000 participants) and the UK Biobank Axiom (∼450,000 participants), which share 95% marker content. Imputed genetic data were obtained by using the Haplotype Reference Consortium (HRC), or the merged 1000 Genomes Project and UK10K as the reference panels.

### Definition of Sex and the Polygenic Risk Score

Sex was determined according to the relative intensity of markers on the Y and X chromosomes [18]. The PRS for CRC was built based on 139 of 140 CRC-related risk variants that were identified in a recent genome-wide association study of CRC risk within individuals of European ancestry [15], and were extracted from the UK Biobank (**Supplementary Table S1;** rs377429877 was not measured and thus was not included in the analysis). The score was calculated by summing the number of risk alleles of the respective variants (0, 1, or 2 copies of the risk allele for genotyped loci; imputed dosages for imputed loci). We also calculated a weighted PRS that summed up risk alleles with weights (log of odds ratio of respective SNP) which was applied in sensitivity analyses.

### Definition of Outcomes

The outcomes of this study included the first diagnosis of CRC and death from CRC (primary cause of death) during the follow-up, coded C18-C20 (malignant neoplasm of colon, rectosigmoid junction or rectum) according to the 10^th^ revision of the International Classification of Diseases (ICD-10). Date of complete follow-up of cancer data was 29 February 2020 for England and Wales, and 31 January 2021 for Scotland. The censoring date for death data was 30 September 2021 for England and Wales and 31 October 2021 for Scotland. Pertinent data were provided by population-based cancer registries and national death registries.

### Statistical Analyses

Among participants aged 40-69 years with imputed genetic data, we only included participants identified as “White British” according to self-reported data and results of principal component analysis of ancestry, since participants in the UK Biobank cohort are predominantly White British, and variants used for building the PRS here were identified in a genome-wide association study of European descents. We excluded those with sex mismatch (difference between self-reported and genetically inferred sex) or sex chromosome aneuploidy, those who had a history of inflammatory bowel disease, CRC or bowel cancer screening, and those with a family history of CRC (defined as father, mother or siblings ever diagnosed with CRC. Codes for inflammatory bowel disease or CRC are presented in **Supplementary Table S2**).

We first described the distribution of sex, age and the PRS in the study population. Cox proportional hazards regression was used to assess the associations of sex and the PRS with the risk of CRC occurrence or mortality. The PRS was categorized according to deciles in the study population, and hazard ratios for PRS deciles were calculated using the middle (5^th^ and 6^th^) deciles as the reference. We examined the proportional hazards assumption by Schoenfeld residuals plots for each covariate and did not observe any relevant deviations.

In order to translate hazard ratios (HRs) for male sex and PRS deciles into how many years of age earlier or later men, and individuals in higher or lower PRS deciles would reach comparable risks as the reference group (women or those in the middle PRS deciles), we applied the concept of risk and rate advancement periods (RAPs). Details of the concept and derivation of RAPs, which are applicable for diseases whose risk monotonically increases with age (such as CRC), have been described in detail elsewhere [16]. Here, point estimates of RAPs were calculated from a multivariable Cox regression model (which included sex, PRS deciles, age in years at assessment center as predictor variable) as ratios of the regression coefficients for sex and age and for PRS group and age, respectively. Derivation of 95% confidence intervals for RAPs was performed as previously described [16].

Risk-adapted starting ages of CRC screening for women and men in the different PRS deciles, denoted SA_w_PRS_ and SA_m_PRS_, were then determined from the combination of the RAPs for sex and PRS deciles as

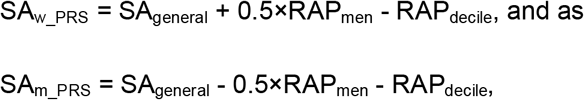

where SA_general_ denotes a general (not risk-adapted) starting age of CRC screening in the population and RAP_men_ and RAP_decile_ denote the RAP estimates for men compared to women and for different PRS deciles compared to the median deciles 5 and 6, respectively.

Exemplary calculations are provided for an overall starting age of 55 years in the general population. From these, the corresponding risk-adapted starting ages for other overall starting ages can be easily derived by adding or subtracting the corresponding difference in the general population starting age.

All statistical analyses were carried out with R software (version 4.2.0).

## Results

### Distribution of age, sex and the PRS in the study population

A total of 242,779 eligible participants were included in this analysis (**Figure 1**), among whom 55.7% were women (**Table 1)**. The proportion of participants in age groups 40-49, 50-59 and 60-69 were 29.1%, 39.1% and 31.7%, respectively. Median (interquartile range) age and PRS were 55 years (48-61) and 134 (129-139) risk alleles. The distributions of age and the PRS were very similar among men and women.

**Table 1.** Characteristics of study population

| **Characteristics** |  | **Both sexes**<br>**(N = 242,779)** | **Women**<br>**(N = 135,161)** | **Men**<br>**(N = 107,618)** |
| --- | --- | --- | --- | --- |
| Sex | Female | 135,161 (55.7) |  |  |
|  | Male | 107,618 (44.3) |  |  |
| Age | 40-49 | 70,751 (29.1) | 39,266 (29.1) | 31,485 (29.3) |
|  | 50-59 | 94,970 (39.1) | 54,162 (40.1) | 40,808 (37.9) |
|  | 60-69 | 77,058 (31.7) | 41,733 (30.9) | 35,325 (32.8) |
|  | Median | 55 | 55 | 55 |
|  | IQR | (48, 61) | (48, 61) | (48, 61) |
| PRS | Median | 134 | 134 | 134 |
|  | IQR | (129, 139) | (129, 139) | (129, 138) |

**Note:** N (%) are presented for categorical variables.
Abbreviations: PRS, polygenic risk score; IQR, interquartile range.

**Figure 1.**
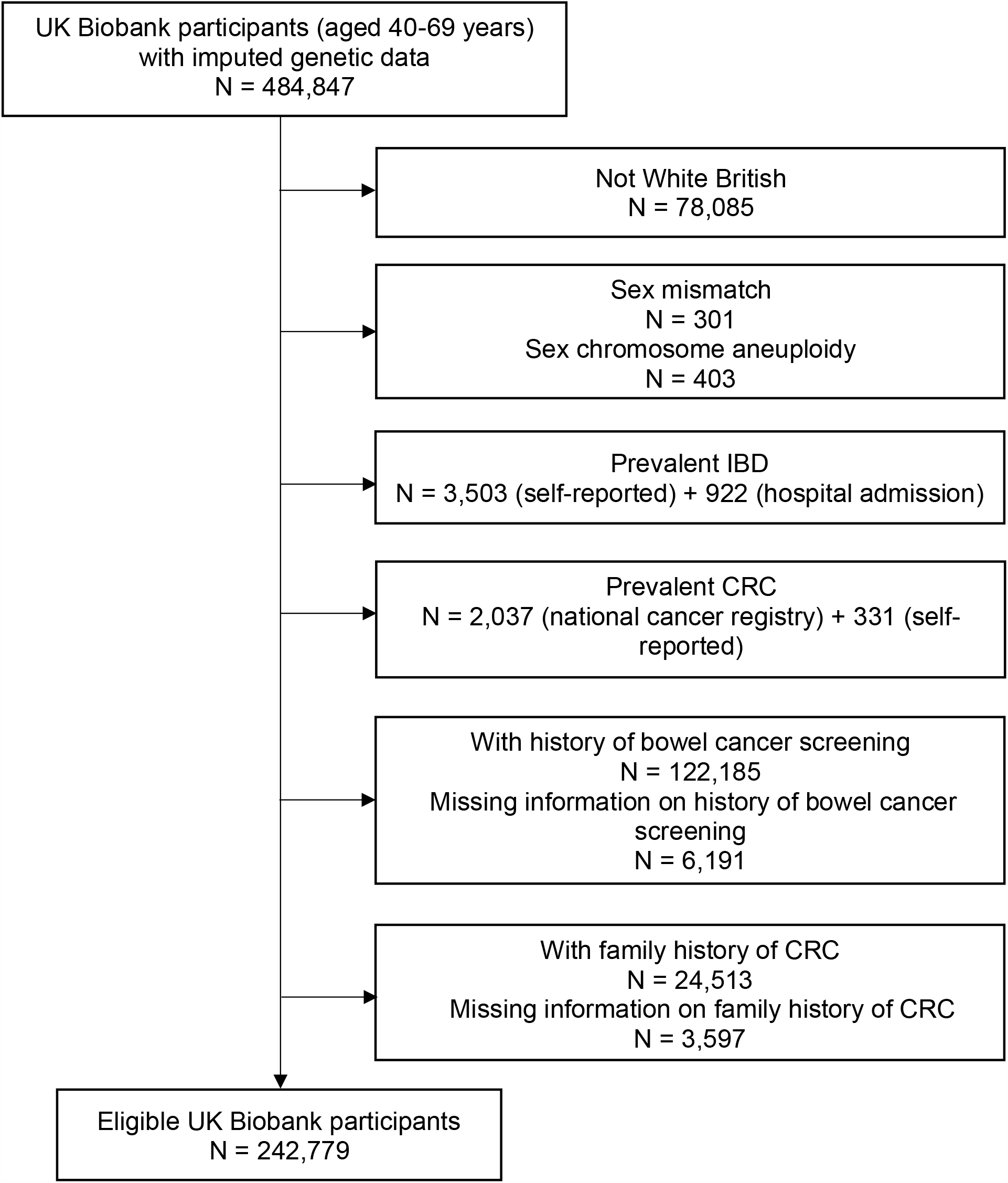
Flow diagram showing selection of study participants from UK Biobank study. Abbreviations: CRC, colorectal cancer; IBD, inflammatory bowel disease.

### Risks and RAPs according to sex and PRS

During a median follow-up time of 11.2 and 12.8 years, 2,714 participants were diagnosed with CRC and 758 died from CRC, respectively. Male sex was associated with a 1.6-fold [Hazard ratio (HR) 1.57, 95% confidence interval (CI) 1.46-1.70] increased risk of CRC occurrence (**Table 2**). Men reached the equivalent CRC risk approximately 6 years (RAP 5.6, 95% CI 4.6-6.6) earlier than women. The PRS was strongly related to CRC risk in a dose-response manner. Compared to those in the middle deciles, people in the lowest and highest PRS decile had approximately half and double the risk (HRs 0.51 and 2.29, respectively), and reached equivalent levels of risk at 8 years older and 10 years younger ages (RAPs -8.4 and 10.3 years, respectively). Similar associations and corresponding RAPs were observed with CRC mortality (RAPs for men vs. women: 4.8 years; for lowest and highest vs middle PRS deciles: -9.3 and 7.9 years, respectively) (**Table 3**). Sensitivity analyses based on a weighted PRS yielded very similar results (**Supplementary Table S3** and **Table S4**).

**Table 2.** Hazard ratios and risk advancement periods regarding the risk CRC occurrence according to sex or polygenic risk score

| **Sex, PRS**<br>**(n risk alleles)** | **N participants** | **N**<br>**CRC cases** | **HR (95% CI)<sup>a</sup>** | **RAP (95% CI)** |
| --- | --- | --- | --- | --- |
| Women | 135,161 | 1,206 | 1.00 (Ref) | 0.0 (Ref) |
| Men | 107,618 | 1,508 | 1.57 (1.46, 1.70) | 5.6 (4.6, 6.6) |
| 1<sup>st</sup> decile ($\leq 124$) | 24,893 | 124 | 0.51 (0.41, 0.62) | -8.4 (-11.0, -5.9) |
| 2<sup>nd</sup> decile (125-128) | 32,858 | 250 | 0.77 (0.66, 0.91) | -3.2 (-5.2, -1.2) |
| 3<sup>rd</sup> decile (129-130) | 22,899 | 175 | 0.77 (0.65, 0.92) | -3.2 (-5.4, -1.0) |
| 4<sup>th</sup> decile (131-132) | 25,640 | 257 | 1.02 (0.87, 1.19) | 0.2 (-1.8, 2.1) |
| 5<sup>th</sup> and 6<sup>th</sup> decile (133-135) | 39,602 | 388 | 1.00 (Ref) | 0.0 (Ref) |
| 7<sup>th</sup> decile (136-137) | 24,831 | 303 | 1.25 (1.08, 1.45) | 2.8 (0.9, 4.6) |
| 8<sup>th</sup> decile (138-140) | 30,413 | 424 | 1.42 (1.24, 1.63) | 4.4 (2.6, 6.1) |
| 9<sup>th</sup> decile (141-143) | 20,466 | 324 | 1.62 (1.40, 1.88) | 6.0 (4.1, 7.8) |
| 10<sup>th</sup> decile ($> 143$) | 21,177 | 469 | 2.29 (2.01, 2.62) | 10.3 (8.5, 12.1) |

<sup>a</sup>Variables in the models included age at attending assessment center, sex and the PRS. Abbreviations: CI, confidence interval; CRC, colorectal cancer; HR, hazard ratio; PRS, polygenic risk score; RAP, risk advancement period; Ref, reference.

**Table 3.** Hazard ratios and risk advancement periods regarding CRC mortality risk according to sex or polygenic risk score

| **Sex, PRS**<br>**(n risk alleles)** | **N participants** | **N**<br>**CRC deaths** | **HR (95% CI)<sup>a</sup>** | **RAP (95% CI)** |
| --- | --- | --- | --- | --- |
| Women | 135,161 | 342 | 1.00 (Ref) | 0.0 (Ref) |
| Men | 107,618 | 416 | 1.53 (1.33, 1.77) | 4.8 (3.1, 6.5) |
| 1<sup>st</sup> decile ($\leq 124$) | 24,893 | 33 | 0.44 (0.30, 0.65) | -9.3 (-13.7, -4.8) |
| 2<sup>nd</sup> decile (125-128) | 32,858 | 68 | 0.69 (0.51, 0.92) | -4.2 (-7.6, -0.9) |
| 3<sup>rd</sup> decile (129-130) | 22,899 | 54 | 0.78 (0.56, 1.07) | -2.8 (-6.4, 0.8) |
| 4<sup>th</sup> decile (131-132) | 25,640 | 62 | 0.80 (0.59, 1.08) | -2.5 (-6.0, 0.9) |
| 5<sup>th</sup> and 6<sup>th</sup> decile (133-135) | 39,602 | 119 | 1.00 (Ref) | 0.0 (Ref) |
| 7<sup>th</sup> decile (136-137) | 24,831 | 79 | 1.06 (0.80, 1.41) | 0.7 (-2.5, 3.9) |
| 8<sup>th</sup> decile (138-140) | 30,413 | 122 | 1.34 (1.04, 1.72) | 3.3 (0.4, 6.1) |
| 9<sup>th</sup> decile (141-143) | 20,466 | 94 | 1.53 (1.17, 2.00) | 4.8 (1.7, 7.9) |
| 10<sup>th</sup> decile ($> 143$) | 21,177 | 127 | 2.02 (1.57, 2.59) | 7.9 (4.9, 10.8) |

<sup>a</sup>Variables in the models included age at attending assessment center, sex and the unweighted PRS.
Abbreviations: CI, confidence interval; CRC, colorectal cancer; HR, hazard ratio; PRS, polygenic risk score; RAP, risk advancement period; Ref, reference.

### Risk-adapted starting ages according to sex and PRS

As an alternative to a general population starting age at age 55, risk-adapted starting screening ages for women would range from 48 years in the highest decile of PRS to 66 years in the lowest decile of PRS (**Table 4**). The corresponding range for men would be from 42 years in the highest PRS decile to 60 years in the lowest PRS decile. Taking CRC mortality as the benchmark, risk-adapted starting ages would range from 50-67 years among women and 45-62 years among men. Risk-adapted starting ages for alternative overall starting ages can be easily derived from the results shown in **Table 4**, e.g. by subtracting or adding 5 years for overall starting ages at 50 or 60 years.

**Table 4.** Exemplary calculation for risk adapted starting age according to sex and PRS decile as an alternative to a general population starting age at age 55

| PRS decile | Risk adapted starting age of screening (years) |  |  |  |  |
| --- | --- | --- | --- | --- | --- |
|  | Risk: CRC incidence |  |  | Risk: CRC mortality |  |
|  | Women | Men |  | Women | Men |
| 1 | 66 | 60 |  | 67 | 62 |
| 2 | 61 | 55 |  | 62 | 57 |
| 3 | 61 | 55 |  | 61 | 56 |
| 4 | 58 | 52 |  | 61 | 56 |
| 5 or 6 | 58 | 52 |  | 58 | 53 |
| 7 | 55 | 49 |  | 57 | 52 |
| 8 | 54 | 48 |  | 55 | 50 |
| 9 | 52 | 46 |  | 53 | 48 |
| 10 | 48 | 42 |  | 50 | 45 |

Abbreviations: CRC, colorectal cancer; PRS, polygenic risk score.

## Discussion

We used the straightforward approach of risk advancement periods to derive risk-adapted starting ages for CRC screening according to sex and a PRS using data from a large UK cohort. Men reached equivalent risk of CRC occurrence and mortality at 5 to 6 years younger ages than women. PRSs were also strongly related to CRC risk and mortality, with people in the highest or lowest PRS decile reaching comparable risk at 8 to 10 years younger or older ages compared to those in the middle PRS deciles, respectively. Based on the combination of genetically defined sex and PRS, risk-adapted starting ages may vary by as much as 24 years between men in the highest PRS decile and women in the lowest PRS decile.

Sex is one of the well-established factors associated with colorectal neoplasms, and is also associated with screening uptake and effectiveness, highlighting the potential importance of sex-specific approaches in prevention and early detection [20,21]. Accumulating evidence has shown several potential mechanisms underlying the impact of sex in the development of CRC [22–25]. Recently, it has been estimated that approximately 50% of the male excess risk of advanced colorectal neoplasm can be explained by known risk factors (e.g., medical, lifestyle and dietary factors) [24]. This high proportion implies that the “unadjusted” sex variable by itself, as applied in our analysis, may reflect the impact of those factors to a substantial degree. Besides, hormonal effects and some unique patterns in gene, protein expression and endocrine cellular signaling in women, were found to contribute to substantially lower CRC risk among women [22,23,25].

The potential relevance of sex differences in CRC risk for starting ages of CRC screening has been outlined in previous studies. Based on cancer registry data from the US in 2000-2003, Brenner et al [26] estimated that women reach risk levels (cumulative 10-year incidence or mortality of CRC) of men aged 50, 55 or 60 years at 4 to 8 years older ages. Similar sex differences in CRC incidence and mortality were also quite consistently found in 38 European countries based on the GLOBOCAN 2002 database [27]. The RAP estimates for sex derived in our study (5.6 and 4.8 years for CRC incidence and mortality, respectively) are in line with these observations.

In the past two decades, large genome-wide association studies have disclosed a rapidly increasing number of single nucleotide polymorphisms (SNPs) that are associated with CRC risk. Although contribution of single SNPs to risk prediction is very small, the combination of a large number of SNPs into a PRS enables quite robust risk discrimination [14,15,28]. However, few studies have attempted to translate differences in PRS into different starting ages of screening.

Jeon et al [7] derived and validated a combined E-score (based on 19 lifestyle and environmental factors) and G-score (based on 63 CRC-related risk variants) using data of multiple case-control and cohort studies from different parts of the world. They combined the risk score with external CRC incidence rates for non-Hispanic whites in the US to derive 10-year absolute risks and risk adapted sex-specific starting ages of CRC screening. Among those with no family history of CRC, these starting ages varied by 12 years between men at the 10^th^ and 90^th^ risk percentile, and by 14 years between women at the 10^th^ and 90^th^ risk percentile. Comparing 1^st^ and 99^th^ percentiles, differences in starting ages were 20 and 25 years, respectively. In addition, for defined risk percentiles, risk-adapted starting ages were between 5 and 10 years lower for men than for women (7 years at the 50^th^ percentile). Using the straightforward approach of RAPs, we derived remarkably consistent differences in starting ages of CRC screening from a single large population-based cohort. Using the results of our analysis, risk-adapted starting ages of screening based on sex and PRS could be derived by a single laboratory analysis without the need of extensive collection of additional risk factor data.

It is worth noting, however, that the RAP approach could be easily extended to derive refined starting ages of screening by running more complex Cox regression models which include additional CRC risk factors. From such models, personalized RAPs could be derived by ratios of risk scores based on the levels and regression coefficients of multiple risk factors including sex and PRS, divided by regression coefficients for age.

However, some issues regarding the implementation of genetics-based prevention approaches merit further consideration. Although there are already commercially available test kits for PRSs [29–31] for several diseases including CRC [30], costs of genetic testing and pre- and post-test clinical support might limit their population-wide application in screening. Genetic testing also raises special ethical issues with respect to confidentiality and privacy protection, not only to patients themselves but also to their biological relatives. Besides, several challenges such as construction of genetic assays that are valid in a diverse population, and interpretation of genetic results to unaffected individuals would need to be addressed before potential clinical implementation.

Research is under way aimed at developing pipelines on how to incorporate information from PRSs in routine clinical practice, including construction of clinically valid assays, interpretation for individual patients, and the development of clinical workflows and resources to support their use in patient care [32]. An ongoing randomized trial aims to assess, if and by much determining a PRS-derived personalized CRC risk estimate in primary care may increase risk-appropriate use of CRC screening [33]. Emerging evidence in this field may help to define the potential role of genetic testing for risk-adapted screening strategies whose pros and cons will have to be weighed against those of other approaches. Latter may include risk stratification by risk factor scores or other biological measurements, such as fecal hemoglobin concentrations below the commonly employed thresholds in CRC screening by fecal immunochemical tests, or combinations of various types of scores [34–37].

A major strength of our study is its reliance on data from a well-designed very large cohort study, the UK Biobank, whose large case numbers and long follow-up enabled deriving risk estimates at very high levels of precision. The parameters needed for the risk estimates were exclusively based on time-invariant biological parameters that can be derived from a single biospecimen, such as saliva or blood. Application of the concept of RAPs enabled derivation of risk-adapted starting ages from the same cohort, without the need to combine cohort data with external data, such as cancer registry data.

There are also some limitations that require careful consideration. First, our analysis was exclusively based on a PRS derived in populations of European ancestry and carried out in a British population of European ancestry. PRSs based on individuals of one genetic ancestry are less predictive in other ancestries [38], which limits generalizability of results. Second, generalizability may further be limited by selective participation of more health-conscious and more healthy people in the UK Biobank [39]. However, this should be of less concern in our analysis which is exclusively based on genetic data than in studies that include health behavior and health outcome data. Third, our analysis was restricted to people without family history of CRC. We deliberately chose to do so as there are established guidelines for earlier starting ages of screening for those with a family history, and as we aimed to focus on time-invariant risk predictors in the average risk population. In contrast to genetically determined sex and PRS, family history of CRC is subject to change during the life course [13]. Also, risk adjusted starting ages according to specific constellations of family history have been examined in great detail in recent studies [9]. Fourth, our analyses are based on relatively simple PRSs based on unweighted or weighted risk allele counts. More sophisticated approaches and inclusion of additional risk variants may further improve PRS-based risk prediction [15,40].

In conclusion, our study demonstrates straightforward derivation of risk-adapted starting ages of CRC screening in the “average risk population” through the estimation of RAPs in the large cohort of the UK Biobank. Our results of a difference of up to 24 years in starting age between men in the highest PRS decile and women in the lowest PRS decile could have far-reaching implications. The RAP approach could be easily extended to incorporate additional risk factor information. Further research should assess feasibility, acceptance and cost-effectiveness of the use of this straightforward approach and alternative risk-adapted approaches to CRC screening.

## Data Availability

This work used data provided by patients and collected by the NHS as part of their care and support. The UK Biobank is an open-access resource and bona fide researchers can apply to use the UK Biobank dataset by registering and applying at https://www.ukbiobank.ac.uk/enable-your-research/apply-for-access. Further information is available from the corresponding author upon request.

## Contributors

Study concept and design: H.B.; Acquisition of data: M.H. and H.B.; Analysis and interpretation of data: X.C., T.H., R.C., M.H., and H.B.; Drafting of the manuscript: X.C. and H.B.; Critical revision of the manuscript for important intellectual consent: X.C., T.H., R.C., M.H., and H.B.; Statistical analysis: X.C., T.H., R.C., M.H., and H.B.; Obtained funding: H.B.; Study supervision: H.B. All authors read and approved the final manuscript.

## Patient and Public Involvement

Patients or the public were not involved in the design, or conduct, or reporting, or dissemination plans of our research.

