## Supplementary Table S1-S4 for "Using risk advancement periods to derive starting ages of colorectal cancer screening according to sex and polygenic risk score: Results from the UK Biobank"

**Supplementary Table S1.** Overview on colorectal cancer related single-nucleotide polymorphisms identified in genome-wide association studies and considered in this analysis

| **SNP** | **Locus** | **Position** | **Risk allele** | **Beta** |
| --- | --- | --- | --- | --- |
| rs4360494 | 1p34.3 | 38455891 | G | 0.0379 |
| rs12144319 | 1p32.3 | 55246035 | C | 0.0661 |
| rs72647484 | 1p36.12 | 22587728 | T | 0.0504 |
| rs7542665 | 1p31.3 | 62673037 | C | 0.0334 |
| rs6678517 | 1q25.3 | 183002639 | A | 0.073 |
| rs17011141 | 1q41 | 222112634 | G | 0.0877 |
| rs448513 | 2q24.2 | 159964552 | C | 0.0054 |
| rs11884596 | 2q33.1 | 199612407 | C | 0.0342 |
| rs983402 | 2q33.1 | 199781586 | T | 0.0622 |
| rs7606562 | 2p16.3 | 48686695 | T | 0.0414 |
| rs11692435 | 2q11.2 | 98275354 | G | 0.0492 |
| rs3731861 | 2q35 | 219191256 | T | 0.0613 |
| rs10049390 | 3q22.2 | 133701119 | A | 0.0455 |
| rs13086367 | 3q13.2 | 112903888 | A | 0.0463 |
| rs72942485 | 3q13.2 | 112999560 | G | 0.0545 |
| rs9831861 | 3p21.1 | 53088285 | G | 0.0294 |
| rs35470271 | 3p22.1 | 40915239 | G | 0.0994 |
| rs12635946 | 3q13.2 | 112916918 | C | 0.0334 |
| rs113569514 | 3q22.2 | 133748789 | T | 0.0414 |
| rs9876206 | 3q26.2 | 169517436 | C | 0.0453 |
| rs6781752 | 3p14.1 | 66365163 | A | 0.0597 |
| rs11727676 | 4q31.21 | 145659064 | C | 0.0093 |
| rs1391441 | 4q24 | 106128760 | A | 0.0148 |
| rs13149359 | 4q22.2 | 94938618 | A | 0.052 |
| rs7708610 | 5p13.1 | 40102443 | A | 0.0384 |
| rs78368589 | 5p15.33 | 1240204 | T | 0.0786 |
| rs145364999 | 5q21.1 | 98206082 | T | 0.3496 |
| rs2735940 | 5p15.33 | 1296486 | G | 0.0865 |
| rs12514517 | 5p13.1 | 40280076 | A | 0.1013 |
| rs755229494 | 5q22.2 | 112097351 | G | 0.6286 |
| rs12659017 | 5q23.2 | 125988175 | G | 0.0374 |
| rs4976270 | 5q31.1 | 134467220 | C | 0.0693 |
| rs13204733 | 6p12.1 | 55566108 | G | 0.0643 |
| rs116685461 | 6p21.33 | 31315512 | G | 0.0655 |
| rs9271695 | 6p21.32 | 32593080 | G | 0.0889 |
| rs2516420 | 6p21.33 | 31449620 | C | 0.1091 |
| rs116353863 | 6p21.33 | 31010185 | C | 0.1202 |
| rs16878812 | 6p21.31 | 35569562 | A | 0.0778 |
| rs9470361 | 6p21.2 | 36623379 | A | 0.054 |
| rs62404966 | 6p12.1 | 55712124 | C | 0.0724 |
| rs3131043 | 6p21.33 | 30758466 | G | 0.0294 |
| rs2070699 | 6p24.1 | 12292772 | T | 0.0294 |
| rs1476570 | 6p22.1 | 29809860 | A | 0.0492 |
| rs3830041 | 6p21.32 | 32191339 | T | 0.0645 |
| rs6928864 | 6q21 | 105966894 | C | 0.0531 |
| rs62396735 | 6p21.1 | 41702582 | C | 0.033 |
| rs12672022 | 7p13 | 45136423 | T | 0.0067 |
| rs80077929 | 7p12.3 | 46094089 | T | 0.0093 |
| rs10951878 | 7p12.3 | 46926695 | C | 0.0531 |
| rs3801081 | 7p12.3 | 47511161 | G | 0.0253 |
| rs7013278 | 8q24.21 | 128414892 | T | 0.0091 |
| rs4313119 | 8q24.21 | 128571855 | G | 0.0518 |
| rs16892766 | 8q23.3 | 117630683 | C | 0.2099 |
| rs6469654 | 8q23.3 | 117632965 | G | 0.0677 |
| rs117079142 | 8q24.11 | 117790914 | A | 0.1139 |
| rs6983267 | 8q24.21 | 128413305 | G | 0.1052 |
| rs34405347 | 9q22.33 | 101679752 | T | 0.0089 |
| rs1537372 | 9p21.3 | 22103183 | G | 0.012 |
| rs10980628 | 9q31.3 | 113671403 | C | 0.0511 |
| rs12217641 | 10p14 | 8663875 | C | 0.0069 |
| rs10786560 | 10q24.2 | 101315166 | G | 0.0082 |
| rs1250567 | 10q22.3 | 81046265 | C | 0.047 |
| rs11255841 | 10p14 | 8739580 | T | 0.1064 |
| rs10821907 | 10q11.23 | 52648454 | C | 0.073 |
| rs704017 | 10q22.3 | 80819132 | G | 0.0765 |
| rs11190164 | 10q24.2 | 101351704 | G | 0.0889 |
| rs12246635 | 10q25.2 | 114288619 | C | 0.0975 |
| rs11196170 | 10q25.2 | 114722621 | A | 0.0527 |
| rs7946853 | 11q13.4 | 74409077 | C | 0.0119 |
| rs55864876 | 11q22.1 | 100717136 | G | 0.015 |
| rs2186607 | 11q22.1 | 101656397 | T | 0.0483 |
| rs61389091 | 11q13.4 | 74427921 | C | 0.1934 |
| rs4450168 | 11p15.4 | 10286755 | C | 0.0413 |
| rs174533 | 11q12.2 | 61549025 | G | 0.0636 |
| rs7121958 | 11q13.4 | 74280012 | G | 0.078 |
| rs3087967 | 11q23.1 | 111156836 | T | 0.1122 |
| rs4759277 | 12q13.3 | 57533690 | A | 0.0285 |
| rs1427760 | 12q24.21 | 115100714 | C | 0.0424 |
| rs3217874 | 12p13.32 | 4400808 | T | 0.0453 |
| rs10849433 | 12p13.31 | 6406904 | C | 0.0468 |
| rs11610543 | 12q12 | 43134191 | G | 0.0474 |
| rs35808169 | 12p13.32 | 4368607 | C | 0.089 |
| rs3217810 | 12p13.32 | 4388271 | T | 0.1181 |
| rs2250430 | 12p13.31 | 6421174 | T | 0.0597 |
| rs77969132 | 12p11.21 | 31594813 | T | 0.1583 |
| rs12372718 | 12q13.12 | 51171090 | G | 0.0896 |
| rs597808 | 12q24.12 | 111973358 | G | 0.0737 |
| rs7300312 | 12q24.21 | 115890922 | C | 0.066 |
| rs2710310 | 12p13.2 | 12035649 | C | 0.0145 |
| rs78341008 | 13q22.1 | 73791554 | C | 0.0109 |
| rs8000189 | 13q34 | 111075881 | T | 0.0473 |
| rs45597035 | 13q22.1 | 73649152 | A | 0.0495 |
| rs1924816 | 13q22.1 | 73997961 | A | 0.0506 |
| rs7333607 | 13q13.3 | 37462010 | G | 0.0758 |
| rs1330889 | 13q22.3 | 78609615 | C | 0.0453 |
| rs1951864 | 14q22.2 | 54369299 | A | 0.0059 |
| rs17094983 | 14q23.1 | 59189361 | G | 0.0062 |
| rs8020436 | 14q23.1 | 59208437 | A | 0.0294 |
| rs35107139 | 14q22.2 | 54419106 | C | 0.0912 |
| rs4901473 | 14q22.2 | 54445157 | G | 0.0465 |
| rs745213 | 15q23 | 68060389 | G | 0.0072 |
| rs12594720 | 15q22.31 | 67007018 | C | 0.0246 |
| rs56324967 | 15q22.33 | 67402824 | C | 0.0689 |
| rs17816465 | 15q13.3 | 33156386 | A | 0.069 |
| rs12708491 | 15q13.3 | 32992836 | G | 0.0464 |
| rs2293581 | 15q13.3 | 33010736 | A | 0.1248 |
| rs7495132 | 15q26.1 | 91172901 | T | 0.0453 |
| rs9930005 | 16q23.2 | 80043258 | C | 0.0061 |
| rs12447408 | 16q24.1 | 86252544 | A | 0.0079 |
| rs9924886 | 16q22.1 | 68743939 | A | 0.055 |
| rs12149163 | 16q24.1 | 86339315 | T | 0.0487 |
| rs62042090 | 16q24.1 | 86703949 | T | 0.0481 |
| rs983318 | 17q24.3 | 70413253 | A | 0.0397 |
| rs73975586 | 17p13.3 | 814243 | A | 0.0497 |
| rs1078643 | 17p12 | 10707241 | A | 0.0747 |
| rs75954926 | 17q25.3 | 81061048 | G | 0.0882 |
| rs373585858 | 17q25.3 | 80394556 | A | 0.1103 |
| rs4968127 | 17p13.3 | 809643 | G | 0.0514 |
| rs11874392 | 18q21.1 | 46453156 | A | 0.1606 |
| rs73068325 | 19q13.43 | 59079096 | T | 0.0066 |
| rs34797592 | 19p13.11 | 16417198 | T | 0.0824 |
| rs28840750 | 19q13.11 | 33519927 | T | 0.1939 |
| rs1963413 | 19q13.2 | 41871573 | A | 0.0441 |
| rs12979278 | 19q13.33 | 49218602 | T | 0.0293 |
| rs2738783 | 20q13.33 | 62308612 | T | 0.006 |
| rs6067417 | 20q13.13 | 48983697 | C | 0.0331 |
| rs6031311 | 20q13.12 | 42666475 | T | 0.0362 |
| rs6091189 | 20q13.13 | 49256285 | T | 0.0549 |
| rs994308 | 20p12.3 | 6603622 | C | 0.0626 |
| rs28488 | 20p12.3 | 6762221 | T | 0.0714 |
| rs556532366 | 20p12.3 | 8568071 | T | 0.0715 |
| rs189583 | 20p12.3 | 6376457 | G | 0.0795 |
| rs4813802 | 20p12.3 | 6699595 | G | 0.0819 |
| rs11087784 | 20p12.3 | 7740976 | G | 0.0874 |
| rs6066825 | 20q13.13 | 47340117 | A | 0.0719 |
| rs6063514 | 20q13.13 | 49055318 | C | 0.0547 |
| rs13831 | 20q13.32 | 57475191 | G | 0.0334 |
| rs1741640 | 20q13.33 | 60932414 | C | 0.1146 |
| rs6058093 | 20q11.22 | 33213196 | C | 0.045 |

Abbreviations: A, adenine; C, cytosine; G, guanine; T, thymine; SNP, single-nucleotide polymorphism.

**Supplementary Table S2.** Codes in UK Biobank study used to identify cases with inflammatory bowel disease or colorectal cancer

| **Categories** | **Field ID in UK Biobank** | **ICD10 codes** | **ICD9 codes** | **Self-reported cancer’s codes** | **Information source** |
| --- | --- | --- | --- | --- | --- |
| Prevalent inflammatory bowel disease | 41270 and 41271 (ICD10)  41280 and 41281 (ICD9)  20002 (self-reported) | Codes start with K50 (CD) or K51 (UC) | Codes start with  555 (CD) or  556 (UC) | 1461 inflammatory bowel disease  1462 Crohn’s disease  1463 ulcerative colitis | Hospital admission/self-reported |
| Prevalent CRC | 40005 and 40006  (ICD10)  40013 (ICD9)  20001 (self-reported) | Codes start with C18, C19 and C20 | Codes start with 153, 1540 and 1541 | 1020, 1022, 1023 | Cancer registry/  Self-reported |
| Incident CRC | 40005 and 40006 (ICD10) | Codes start with C18, C19 and C20 |  |  | Cancer registry |

Abbreviations: CD, Crohn’s disease; ICD, International Classification of Diseases; UC, ulcerative colitis.

**Supplementary Table S3.** Hazard ratios and risk advancement periods regarding the risk CRC occurrence according to sex or weighted polygenic risk score

| **Sex,**  **weighted PRS** | **N participants** | **N**  **CRC cases** | **HR (95% CI)^a^** | **RAP (95% CI)** |
| --- | --- | --- | --- | --- |
| Women | 135,161 | 1,206 | 1.00 (Ref) | 0.0 (Ref) |
| Men | 107,618 | 1,508 | 1.57 (1.46, 1.70) | 5.6 (4.6, 6.6) |
| 1^st^ decile (≤7.5) | 22,687 | 102 | 0.42 (0.34, 0.52) | -10.8 (-13.5, -8.0) |
| 2^nd^ decile (7.6-7.7) | 24,145 | 154 | 0.60 (0.50, 0.72) | -6.4 (-8.7, -4.1) |
| 3^rd^ decile (7.8-7.9) | 35,761 | 288 | 0.76 (0.65, 0.88) | -3.4 (-5.3, -1.6) |
| 4^th^ decile (8.0) | 20,929 | 203 | 0.91 (0.77, 1.08) | -1.1 (-3.2, 0.9) |
| 5^th^ and 6^th^ decile (8.1-8.2) | 42,915 | 456 | 1.00 (Ref) | 0.0 (Ref) |
| 7^th^ decile (8.3) | 20,243 | 260 | 1.21 (1.04, 1.41) | 2.4 (0.5, 4.3) |
| 8^th^ decile (8.4-8.5) | 33,389 | 397 | 1.13 (0.98, 1.29) | 1.5 (-0.2, 3.1) |
| 9^th^ decile (8.6-8.7) | 22,161 | 372 | 1.59 (1.39, 1.83) | 5.8 (4.0, 7.5) |
| 10^th^ decile (>8.8) | 20,549 | 482 | 2.25 (1.98, 2.56) | 10.0 (8.3, 11.7) |

^a^Variables in the models included age at attending assessment center, sex and the weighted PRS.

Abbreviations: CI, confidence interval; CRC, colorectal cancer; HR, hazard ratio; PRS, polygenic risk score; RAP, risk advancement period; Ref, reference.

**Supplementary Table S4.** Hazard ratios and risk advancement periods regarding CRC mortality risk according to sex or weighted polygenic risk score

| **Sex, weighted**  **PRS** | **N participants** | **N**  **CRC deaths** | **HR (95% CI)^a^** | **RAP (95% CI)** |
| --- | --- | --- | --- | --- |
| Women | 135,161 | 342 | 1.00 (Ref) | 0.0 (Ref) |
| Men | 107,618 | 416 | 1.54 (1.33, 1.77) | 4.8 (3.1, 6.5) |
| 1^st^ decile (≤7.5) | 22,687 | 29 | 0.40 (0.27, 0.59) | -10.4 (-15.1, -5.7) |
| 2^nd^ decile (7.6-7.7) | 24,145 | 46 | 0.59 (0.43, 0.83) | -5.9 (-9.7, -2.0) |
| 3^rd^ decile (7.8-7.9) | 35,761 | 82 | 0.72 (0.55, 0.95) | -3.7 (-6.8, -0.6) |
| 4^th^ decile (8.0) | 20,929 | 52 | 0.78 (0.57, 1.07) | -2.8 (-6.4, 0.8) |
| 5^th^ and 6^th^ decile (8.1-8.2) | 42,915 | 137 | 1.00 (Ref) | 0.0 (Ref) |
| 7^th^ decile (8.3) | 20,243 | 66 | 1.03 (0.76, 1.37) | 0.3 (-3.0, 3.6) |
| 8^th^ decile (8.4-8.5) | 33,389 | 103 | 0.97 (0.75, 1.25) | -0.3 (-3.2, 2.6) |
| 9^th^ decile (8.6-8.7) | 22,161 | 101 | 1.44 (1.11, 1.86) | 4.1 (1.2, 7.0) |
| 10^th^ decile (>8.8) | 20,549 | 142 | 2.19 (1.73, 2.77) | 8.8 (6.0, 11.7) |

^a^Variables in the models included age at attending assessment center, sex and the weighted PRS.

Abbreviations: CI, confidence interval; CRC, colorectal cancer; HR, hazard ratio; PRS, polygenic risk score; RAP, risk advancement period; Ref, reference.
